# Predictors of statin adherence among statin users in primary care using real-world data

**DOI:** 10.64898/2026.02.24.26347032

**Authors:** Shagoofa Rakhshanda, Kerry-Anne Rye, Siaw-Teng Liaw, Joel Rhee, Jitendra Jonnagaddala

**Affiliations:** School of Population Health, UNSW Sydney, NSW 2052 Australia; School of Biomedical Sciences, UNSW Sydney, NSW 2052 Australia; Discipline of General Practice, School of Clinical Medicine, UNSW Sydney, NSW 2052 Australia; SREDH Consortium, Sydney, Australia

## Abstract

**Purpose:** This study aimed to identify the demographic and clinical predictors of statin adherence among patients prescribed statins for the primary prevention of CVD during the first two years following their first recorded statin prescription, using real-world data.

**Methods:** The Electronic Practice Based Research Network Linked Dataset was used in this study. Statin adherence was calculated using a modified proportion of days covered (PDC), based on prescribing data from the date of the first recorded statin prescription to the to the last recorded visit within the two years observation period. Patients with a PDC ≥ 80% were classified as adherent. All analyses were performed using R software. Descriptive analyses and multivariate logistic regression were performed. Backward stepwise selection guided by the Akaike Information Criterion was used to derive a parsimonious final model.

**Results:** Overall, 3,432 patients accounting for 57,227 visits were included. The mean PDC was 91.6% (±22.2%), and 72.0% of patients were classified as adherent. Higher odds of adherence was observed among patients aged over 65 years (adjusted odds ratio [AOR] 1.4, 95% confidence interval [CI] 1.1 – 1.8), those residing in Socio-Economic Indexes for Australia 2021 Index of Relative Socio-Economic Advantage and Disadvantage category 3 (AOR 2.1, 95% CI 1.6 – 2.9), those prescribed three medications, including a statin (AOR 1.8, 95% CI 1.3 – 2.4) and those with one selected comorbidity (AOR 1.4, 95% CI 1.1 – 1.7), compared with their respective reference categories. Patients prescribed two or more statin types had lower odds of adherence (AOR 0.7, 95% CI 0.5 – 0.9).

**Conclusions:** Age, area level socioeconomic status, and the number of prescribed medications, selected comorbidities and statin types were associated with statin adherence. However, adherence was estimated from prescribing rather than dispensing data, and the model demonstrated modest discrimination. Further validation in independent populations is needed to validate these associations.

## 1. Introduction

Nearly one in six Australians live with cardiovascular diseases (CVDs), that are responsible for over 1,500 hospitalisations per year (1). In 2022, CVDs contributed to about 12% of the total burden of disease in Australia, and over 57,000 acute coronary events, such as angina or heart attack, were reported. Moreover, about 24% of all deaths in 2022 were caused by CVD (1, 2). Statins, a group of lipid-modifying drugs, are widely prescribed as primary and secondary prevention of CVD in Australia (3). The 2023 Australian Guideline for assessing and managing CVD risk recommends that healthcare providers prescribe statins to anyone with ≥10% estimated 5-years CVD risk, and to consider prescribing a statin to patients with 5% to <10% risk. A statin is also recommended for patients with total cholesterol >7.5 mmol/L (1). The predictors for statin prescriptions in Australia include age, education, and the presence of comorbidities (4).

Statin adherence is essential for reducing the risk of CVD and preventing adverse health outcomes. Statins can significantly reduce risk of cardiovascular events by reducing plasma cholesterols levels. However, up to 30% of patients that are prescribed a statin are not adherent (5). A previous systematic review found that some studies reported statin adherence rates, across clinical settings, at 20% to 60% (6). Statin adherence is higher for secondary prevention of CVD compared to primary prevention (7, 8). Inconsistent adherence was found to be lower than the recommended threshold among acute coronary syndrome (ACS) patients (9). Lower adherence to statins has also been associated with higher morbidity and mortality, specifically in CVD patients (10, 11).

Several demographic factors contribute to low adherence to statin therapy. Younger patients, especially those aged less than 65 years, exhibit poorer statin adherence than those aged 65 years or above. This may be because older adults consider themselves to be at a higher risk of CV events, which drives them to take prescribed medications more regularly, than younger patients (7, 12, 13, 14). Gender is also a predictor of statin adherence, with females showing poorer adherence that males (7, 15, 16). However, gender was not found to be a predictor of statin adherence in other studies (17, 18). Unemployed people are also less likely to be adherent to statins than their employed counterparts (19, 20, 21, 22, 23). However, some studies contradict this (7), which highlights the necessity for further studies on this topic.

Statin adherence is also poorer among patients without comorbidities, such as diabetes or hypertension, than those with comorbidities that do not have to manage multiple medical conditions, which makes them less vigilant about treatment regimens (24, 25). Smoking and alcohol consumption have been associated with poorer adherence to statins (7, 26). This could be due to psychological aspects, such as smokers who justify their habit by considering themselves at low risk of CV events, or physiological aspects, such as interactions between alcohol and statin drugs which may cause adverse reactions among drinkers (16, 27).

Ethnicity as well as socioeconomic factors are also predictors for statin adherence. For example, statin adherence is lower in minorities including African Americans and Asians (28, 29, 30). Barriers to statin adherence include the economic burden of out-of-pocket costs and lower co-payments (7, 31, 32, 33, 34). However, this is a vicious cycle whereby higher economic burden leads to poorer statin adherence, which in turn contributes to increased CVD risk, disease progression and hospitalizations, leading to higher healthcare costs (9, 35). High intensity statins have also been associated with lower statin adherence, possibly due to increased side effects (36, 37, 38, 39). Conversely, moderate intensity statins are associated with higher statin adherence (37, 40). In light of the diverse factors that influence statin adherence, the objective of this study was to identify the demographic and clinical predictors of statin adherence among patients prescribed statins for the primary prevention of CVD during the first two years following their first recorded statin prescription, using routinely collected general practice EHR data, with a view to generating insights for improving adherence across all groups.

## 2. Methods

### 2.1 Data source

The Electronic Practice Based Research Network (ePBRN) Linked Dataset, a comprehensive electronic health records (EHR) derived from general practice facilities in the Fairfield and Wollondilly regions of the South-west Sydney Local Health District (SWS LHD) was used in this study. This dataset consists of about 249,345 patients spread across 11 general practice sites, who had at least one visit between 2012 and 2019 (41, 42).

The ePBRN dataset consists of 76 tables, 39 of which were sourced from Best Practice, and 37 tables were sourced from Medical Director. A meticulous selection of data from these tables was employed to ensure enhanced precision and relevance. The research objective of this study requires the extraction of data on statin adherence. This is associated with information related to patient diagnosis, conditions, visit details, and demographics, which could be extracted from six tables (Careplan Goal, Current Rx, Visit Reason, Visits, Past History, and Patients) from Best Practice and 5 tables (Diagnosis, Prescription, History, Consultations, and Patients) from Medical Directors.

### 2.2 Selection criteria

We included patients who were prescribed any type of statin found in the ePBRN dataset (atorvastatin, rosuvastatin, simvastatin, pravastatin, or fluvastatin), under any brand name. The list of statin brand names included in the ePBRN dataset is provided in **Supplement 1**.

A Master table containing all patients with a recorded statin prescription was created for this study. For each patient, the date of their earliest statin prescription recorded in the ePBRN dataset was assigned as the index date. The complete EHR history available before this date was examined for evidence of earlier recorded statin use, and patients with a statin prescription identified during this period were excluded. As the duration of available EHR history differed between patients no fixed lookback period was applied. Consequently, the index date indicates the first statin prescription identified within the available ePBRN records and cannot necessarily be considered the patient’s first ever exposure to statin therapy.

Eligibility was restricted to adults aged 18 to 80 years at the index date. Patients were also required to satisfy the RACGP definition of an active patients having at least three general practice visits for any reason during the two years observation period (43). Patients were not eligible if gender or year of birth was not recorded, if they did not survive during the two years following the index date, or if a CVD event was recorded either before the index date or during the subsequent observation period.

The age criteria were applied to define a population appropriate for the study of statin use in primary prevention. Adults from 18 years of age were eligible because statin therapy may be considered for younger adults with high cardiovascular risk (44). In contrast, patients aged over 80 years were excluded because treatment decisions and patterns of statin use may be more complex and variable in advanced ages (45, 46). Patients who did not survive during the observation period were excluded because complete observation over the required two years was not available. Similarly, the exclusion of patients with a CVD event before or during the observation period was because the occurrence of CVD changes the indication for statin therapy and may alter subsequent prescribing and treatment patterns.

For the analysis of predictors of statin adherence, patients with a calculated proportion of days covered (PDC) of 130% or greater were also excluded. These extreme PDC values typically arise from factors such as early refilling, dose changes, or medication stockpiling and may not represent actual medication use. The study eligibility is illustrated in **Figure 1**.

**Figure 1:**
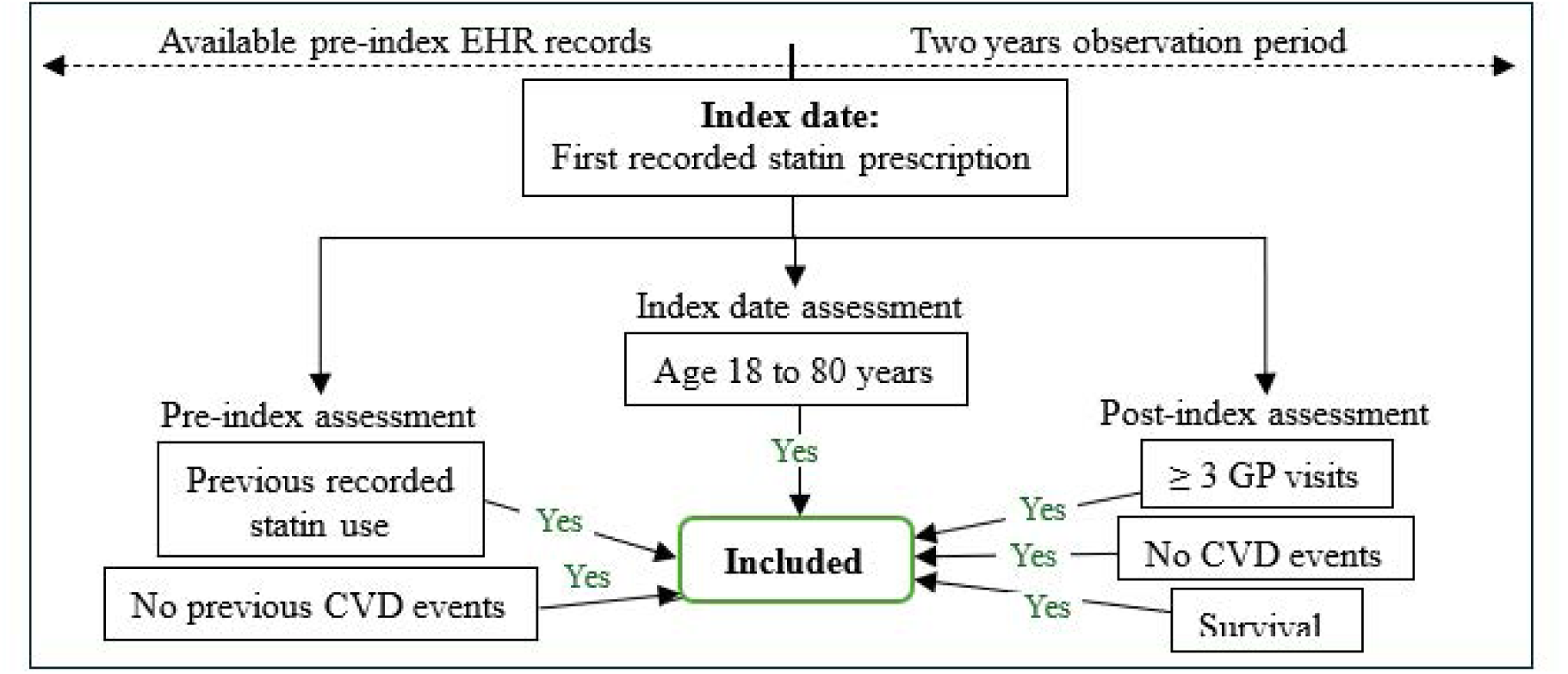
Study eligibility and observation period for the analysis. **Notes:** EHR: Electronic Health Record; GP: General practitioner; CVD: Cardiovascular disease

### 2.3 Outcome

After identifying the patients who met the selection criteria, we calculated 2-years statin adherence using a modified PDC. Conventionally, PDC is calculated as the number of days covered by medication during a defined observation period divided by the total number of days in that observation period. In this study, a modified PDC was used because the ePBRN dataset contains prescription records rather than pharmacy dispensation records, and medication coverage could therefore be estimated from prescribing information but not confirmed from dispensing records. As such, complete medication coverage could not be established for an exact two years period for every patient. For this reason, the maximum PDC was capped greater than 100%.

To ensure that medication coverage was estimated only within the predefined observation period and was not extended beyond two years, the denominator was calculated as the total number of days between the index date and the last recorded GP visit occurring within the two years observation period. Although all included patient ha at least two years of available EHR follow-up after the index date, the timing of their last recorded visit within this period varied and could occur before day 730. The denominator therefore reflected the period over which medication coverage could be estimated from the available prescribing information rather than automatically extending the calculation to day 730 when the last recorded encounter occurred earlier.

The numerator was estimated using prescription information on the number of repeats (the number of repeats of each prescription), quantity (the number of units of the prescribed statin supplied per dispensing, such as tablets or capsules), and frequency (the number of times per day the patient is meant to take the prescribed medication).

For each prescription, the number of authorized supplies was calculated by adding the original supply to the number of repeats recorded in the ePBRN dataset. Accordingly, one was added to the recorded number of repeats before calculating medication coverage. For example, a prescription with five repeats was counted as six authorized supplies, while a prescription without repeats was counted as one authorized supply. For the final prescription interval, information from the second last statin prescription was used to estimate medication coverage up to the last recorded visit, based on the assumption that the prescribed medication could provide coverage during this interval. This approach allowed medication coverage to be estimated within the period defined by the index date and the last recorded visit, without extending the calculation beyond that visit.

The formula used to calculate the numerator was:

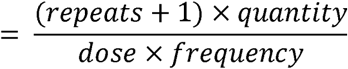

The combined formula for calculating the PDC was:

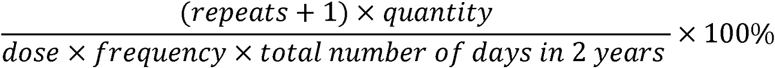

Individuals with PDC ≥ 80% during the two years of observation period were considered adherent (47, 48), and those with PDC < 80% during the observation period were considered non-adherent (48, 49).

### 2.4 Covariates

Patients who were adherent or non-adherent were characterized by age (<30, 30-39, 40-49, 50-59, 60-69 and ≥70), gender, ethnicity, area-based social advantage and disadvantage index, employment status, smoking status, statin intensity at index date (classified as per **Supplement 2**), statin type at index date, number of statin types during the first two years, change in statin intensity between the index date and last visit in two years, number of prescribed medications and number of selected comorbidities. Area-based social advantage and disadvantage was classified using the Socio-Economic Indexes for Australia (SEIFA) 2021 Index of Relative Socio-Economic Advantage and Disadvantage (IRSAD) which is based on suburbs and localities (50).

The number of prescribed medications included the prescribed statin together with any other medications recorded for the patients. Thus, a count of one indicated that only the statin was prescribed, whereas a count of two indicated the statin plus one additional medication. Medications other than statins were classified into anti-diabetic, anti-hypertensive, anti-coagulant, and other medication groups.

The number of selected comorbidities was assessed using a predefined set of conditions available for this analysis: diabetes, hypertension, chronic respiratory disease, cancer, stroke, blood clots, and Parkinson’s disease. The comorbidity count reflected how many of these selected conditions were recorded for each patient rather than the patient’s total number of comorbid conditions. A count of zero indicated that none of the selected conditions were recorded, while a count of one indicated the presence of one condition from the predefined categories.

### 2.5 Statistical analyses

Descriptive analyses were initially conducted to summarise statin adherence according to the covariates and potential predictor variables. Predictors of statin adherence were examined using multivariate logistic regression analysis with backward stepwise model selection approach (48, 51). The initial model included all variables identified a priori as potentially relevant predictors, regardless of their associations in the univariate analyses or the amount of missing information. To account for the incomplete or missing information, multiple imputation by chained equations (MICE) was undertaken (52, 53). Entries classified as “not recorded” were considered missing, and 50 completed datasets were generated using information from the other predictor variables and the statin adherence outcome. The imputation procedure was conducted under the missing at random assumption, whereby missingness was assumed to depend on the observed information included in the imputation model. This method enabled the inclusion of patients with incomplete records in the regression analysis, thereby reducing the loss of observations associated with a complete case approach.

Backward elimination was guided by the Akaike Information Criterion (AIC), with variables removed according to their contribution to the model fit and the model with the lower AIC preferred at each stage (54, 55, 56). The process was used to derive a more parsimonious final model. Backward stepwise model selection was conducted independently for each of the 50 imputed datasets. The model specification that occurred most frequently across these analyses was identified as the final reduced model and subsequently fitted to all imputed datasets. The resulting regression coefficients were then combined according to Rubin’s rules to estimate pooled odds ratios (ORs), 95% confidence intervals (CIs), and p-values. The stability of predictor selection was further assessed using 100 bootstrap resamples of the original dataset with multiple imputation. The assumptions of regression were assessed, and associations between the selected predictors and statin adherence were reported as adjusted ORs (AORs) with 95% CIs. Statistical analyses were conducted using R software version 4.3.2.

This study and the use of data from the ePBRN were approved by the UNSW Human Research Ethics Advisory Panel (HC230066; 23 June 2023). The study was reported in accordance with the European Society for Patient Adherence, COMpliance, and Persistence (ESPACOMP) Medication Adherence Reporting Guideline (EMERGE) guideline for reporting studies of medication adherence, with the completed checklist provided in **Supplement 3**.

## 3. Results

### 3.1 Patient selection

In the ePBRN dataset, 18,377 patients accounting for 110,147 visits were prescribed statins, drawn from a total cohort of 249,345 patients with 3,103,072 visits. After removing patients based on the exclusion criteria (14,945 patients with 52,920 visits), 3,432 patients with 57,227 visits were included in the analysis. The details of patient selection are shown in **Figure 2**.

**Figure 2:**
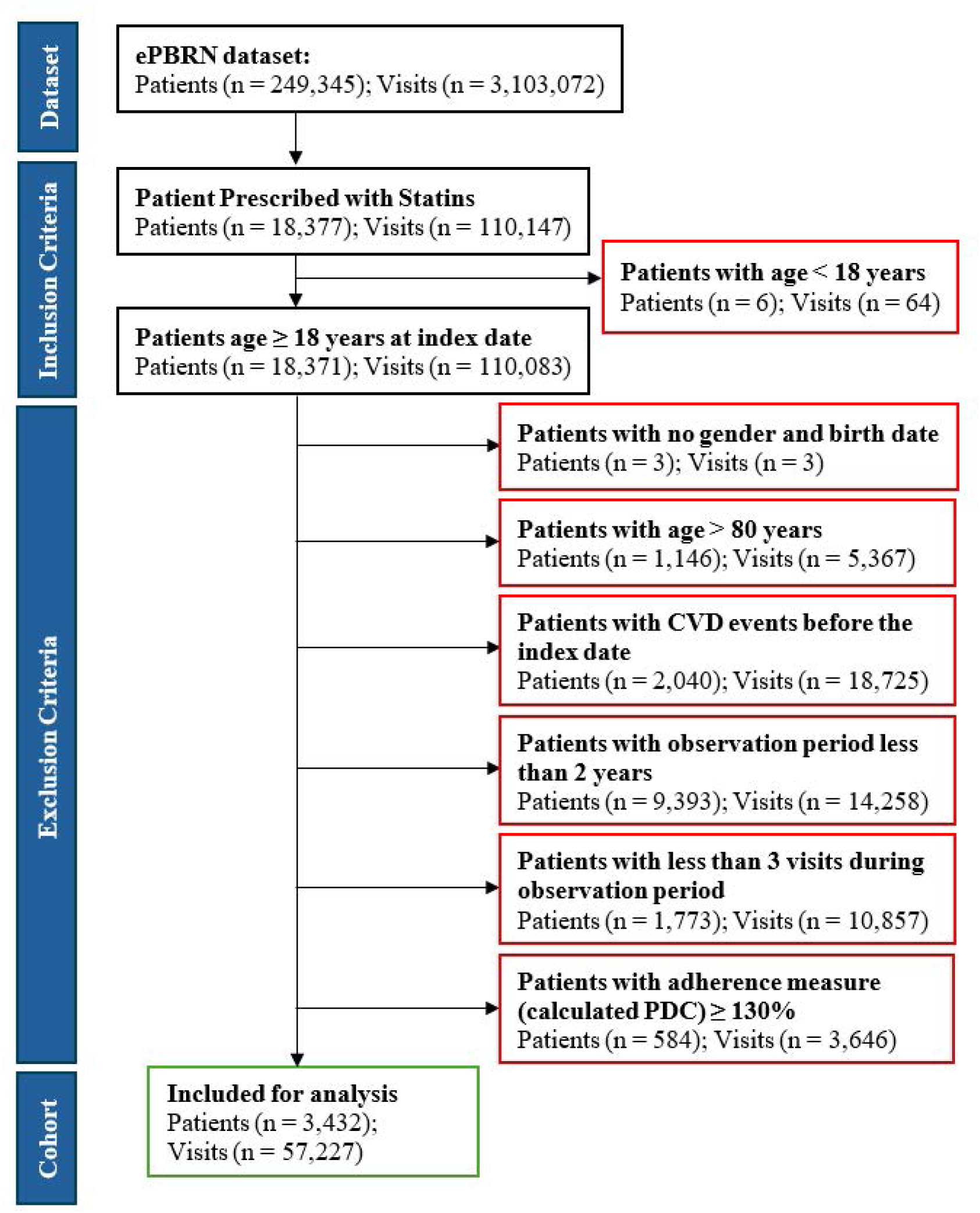
Selection of eligible patients. **Notes:** ePBRN: Electronic Practice Based Research Network; CVD: Cardiovascular disease

### 3.2 Distribution of statin adherence

The distribution of statin adherence during the 2 years of observation period is shown in **Figure 3**. Overall, 2,471 patients (72.0%) were adherent (PDC ≥ 80%) in the first two years after the index date. The mean PDC was 91.6% (±22.2%).

**Figure 3:**
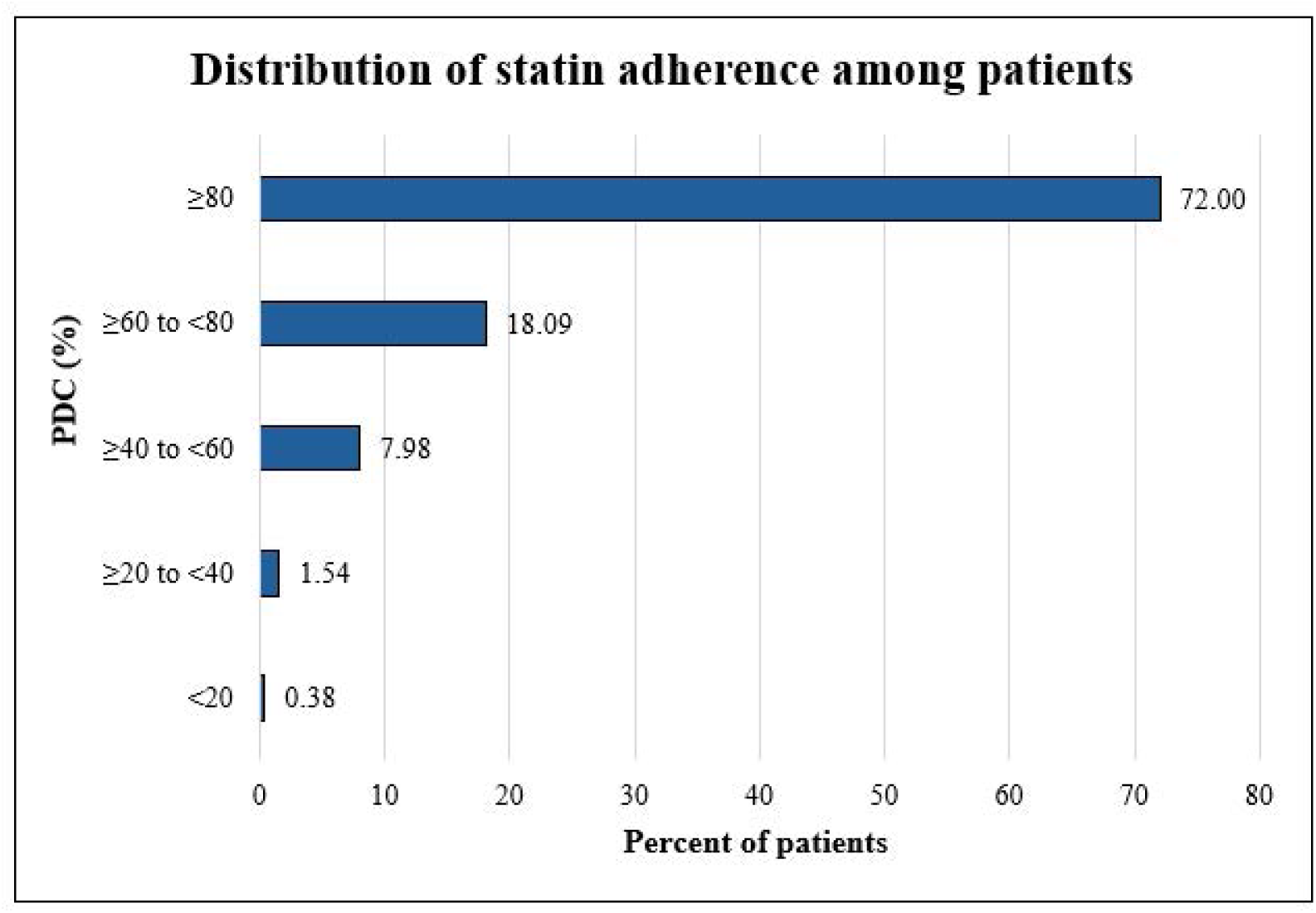
Distribution of statin adherence among patients in first two years after the index date (N= 3,432) **Note:** PDC: Proportion of days covered

### 3.3 Socio-demographic and health-related characteristics of patients

Among the adherent patients, the two most prevalent age groups were 60–69 years (32.2%) and 50–59 years (30.2%) and **(Table 1)**. Approximately half of the adherent patients were males (50.3%). About 75.5% of the adherent patients were recorded as non-Aboriginals and/or Torres Strait Islanders, and 30.4% resided in SEIFA IRSAD category 8 or 9 (representing a relatively socio-economically advantageous population). Employment and smoking status were not recorded for 62.9% and 10.7% of adherent patients, respectively. Among the adherent patients with recorded employment status, 15.7% were employed, while among those with recorded smoking status, 48.0% were non-smokers.

**Table 1:** Characteristics of patients, stratified by adherence status (N = 3,432)

| Variable | Total<br>(N = 3,432) | Adherence status |  | P-value |
| --- | --- | --- | --- | --- |
|  |  | Adherent<br>(n = 2,471) | Non-adherent<br>(n = 961) |  |
| Socio-economic and demographic factors |  |  |  |  |
| Age in years [mean (SD)] | 58.4 (10.9) | 59.4 (10.8) | 55.9 (10.9) | < 0.01 |
| Age in years [n (%)] |  |  |  |  |
| < 30 | 20 (0.58) | 12 (0.49) | 8 (0.83) | < 0.01 |
| 30 – 39 | 148 (4.31) | 81 (3.28) | 67 (6.97) |  |
| 40 – 49 | 542 (15.79) | 360 (14.57) | 182 (18.94) |  |
| 50 – 59 | 1,089 (31.73) | 747 (30.23) | 342 (35.59) |  |
| 60 – 69 | 1,043 (30.39) | 795 (32.17) | 248 (25.81) |  |
| ≥ 70 | 590 (17.19) | 476 (19.26) | 114 (11.86) |  |
| Gender |  |  |  |  |
| Male | 1,731 (50.44) | 1,244 (50.34) | 487 (50.68) | 0.963 |
| Female or other | 1,701 (49.56) | 1,227 (49.66) | 474 (49.32) |  |
| Ethnicity |  |  |  |  |
| Non-ATSI | 2,621 (76.37) | 1,866 (75.52) | 755 (78.56) | < 0.01 |
| ATSI | 29 (0.84) | 17 (0.69) | 12 (1.25) |  |
| Not Recorded <sup>1</sup> | 782 (22.79) | 588 (23.80) | 194 (20.19) |  |
| SEIFA IRSAD category (decile) |  |  |  |  |
| 1 | 595 (17.34) | 428 (17.32) | 167 (17.38) | < 0.01 |
| 2 | 361 (10.52) | 221 (8.94) | 140 (14.57) |  |
| 3 | 501 (14.60) | 395 (15.99) | 106 (11.03) |  |
| 4 | 924 (26.92) | 676 (27.36) | 248 (25.81) |  |
| 8 or 9 | 1,051 (30.62) | 751 (30.39) | 300 (31.22) |  |
| Employment status |  |  |  |  |
| Unemployed | 234 (6.82) | 159 (6.43) | 75 (7.80) | < 0.01 |
| Employed | 616 (17.95) | 389 (15.74) | 227 (23.62) |  |
| Retired and/or pensioner | 466 (13.58) | 370 (14.97) | 96 (9.99) |  |
| Not recorded <sup>1</sup> | 2,116 (61.66) | 1,553 (62.85) | 563 (58.58) |  |
| Health related factors |  |  |  |  |
| Smoking status |  |  |  |  |
| Non-smoker | 1,641 (47.81) | 1,186 (48.00) | 455 (47.35) | < 0.05 |
| Smoker | 1,387 (40.41) | 1,016 (41.12) | 371 (38.61) |  |
| Ex-smoker | 10 (0.29) | 5 (0.20) | 5 (0.52) |  |
| Not recorded <sup>1</sup> | 394 (11.48) | 264 (10.68) | 130 (13.53) |  |

| Statin-related factors |  |  |  |  |
| --- | --- | --- | --- | --- |
| Statin intensity at index date |  |  |  |  |
| High | 732 (21.33) | 514 (20.80) | 218 (22.68) | 0.345 |
| Moderate | 2,523 (73.51) | 1,824 (73.82) | 699 (72.74) |  |
| Low | 177 (5.16) | 133 (5.38) | 44 (4.58) |  |
| Statin type at index date |  |  |  |  |
| Atorvastatin | 1,592 (46.39) | 1,127 (45.61) | 465 (48.39) | 0.232 |
| Rosuvastatin | 1,106 (32.23) | 796 (32.21) | 310 (32.26) |  |
| Simvastatin | 573 (16.70) | 423 (17.12) | 150 (15.61) |  |
| Fluvastatin or Pravastatin | 161 (4.69) | 125 (5.06) | 36 (3.75) |  |
| Number of statin types |  |  |  |  |
| 1 | 3,105 (90.47) | 2,260 (91.46) | 845 (87.93) | < 0.01 |
| ≥ 2 | 327 (9.53) | 203 (8.54) | 116 (12.07) |  |
| Change in statin intensity |  |  |  |  |
| No change | 3,062 (89.22) | 2,201 (89.07) | 861 (89.59) | 0.258 |
| High to low or moderate | 80 (2.33) | 52 (2.10) | 28 (2.91) |  |
| Moderate to high | 219 (6.38) | 163 (6.60) | 56 (5.83) |  |
| Moderate to low | 20 (0.58) | 13 (0.53) | 7 (0.73) |  |
| Low to moderate or high | 51 (1.49) | 42 (1.70) | 9 (0.94) |  |
| Number of prescribed medications <sup>2</sup> |  |  |  |  |
| 1 <sup>3</sup> | 230 (6.70) | 141 (5.71) | 89 (9.26) | < 0.01 |
| 2 | 2,272 (66.20) | 1,595 (64.55) | 677 (70.45) |  |
| 3 | 818 (23.83) | 645 (26.10) | 173 (18.00) |  |
| ≥ 4 | 112 (3.26) | 90 (3.64) | 22 (2.29) |  |
| Number of selected comorbidities <sup>4</sup> |  |  |  |  |
| None | 2,196 (63.99) | 1,506 (60.95) | 690 (71.80) | < 0.01 |
| 1 | 936 (27.27) | 724 (29.30) | 212 (22.06) |  |
| 2 | 277 (8.07) | 224 (9.07) | 53 (5.52) |  |
| ≥ 3 | 23 (0.67) | 17 (0.69) | 6 (0.62) |  |

**Notes:**
<sup>1</sup> Not recorded categories represent missing EHR information and should not be interpreted as patient-level clinical predictors;
<sup>2</sup> The categories for prescribed medications included anti-diabetic drugs, anti-hypertensive drugs, anti-coagulant drugs and one or more of other drugs;
<sup>3</sup> The number of prescribed medications = 1 are the people who are only on a statins;
<sup>4</sup> The categories for comorbidities included diabetes, hypertension, chronic respiratory disease, cancer, stroke, blood clots and Parkinson's disease;
ATSI: Aboriginals and Torres Strait Islanders; SEIFA IRSAD: Socio-Economic Indexes for Areas Index of Relative Socio-Economic Advantage and Disadvantage (with lower categories indicating greater disadvantage and higher categories indicating greater advantage)

At the index date, 45.6% of the adherent patients were prescribed atorvastatin, and 73.8% were prescribed a moderate intensity statin. During the observation period, 91.5% patients had no change in the statin type and 89.1% patients had no change in statin intensity. Approximately 64.6% of the adherent patients had two prescribed medications recorded, representing the statin and one additional medication. In relation to the selected comorbidities included in the analysis, 61.0% of patients had none of these conditions recorded.

### 3.4 Factors affecting statin adherence

**Table 2** presents the univariate and multivariate logistic regression analyses of factors associated with statin adherence. After adjusting for all the variables included in the model, higher odds of statin adherence were observed among patients aged over 65 years (AOR 1.4, 95% CI 1.1 – 1.8), those residing in SEIFA IRSAD category 3 (AOR 2.1, 95% CI 1.6 – 2.9), those prescribed three medications, including a statin and two other medications (AOR 1.8, 95% CI 1.3 – 2.4), and those with one selected comorbidity (AOR 1.4, 95% CI 1.1 – 1.7), compared with their respective reference categories.

**Table 2:**
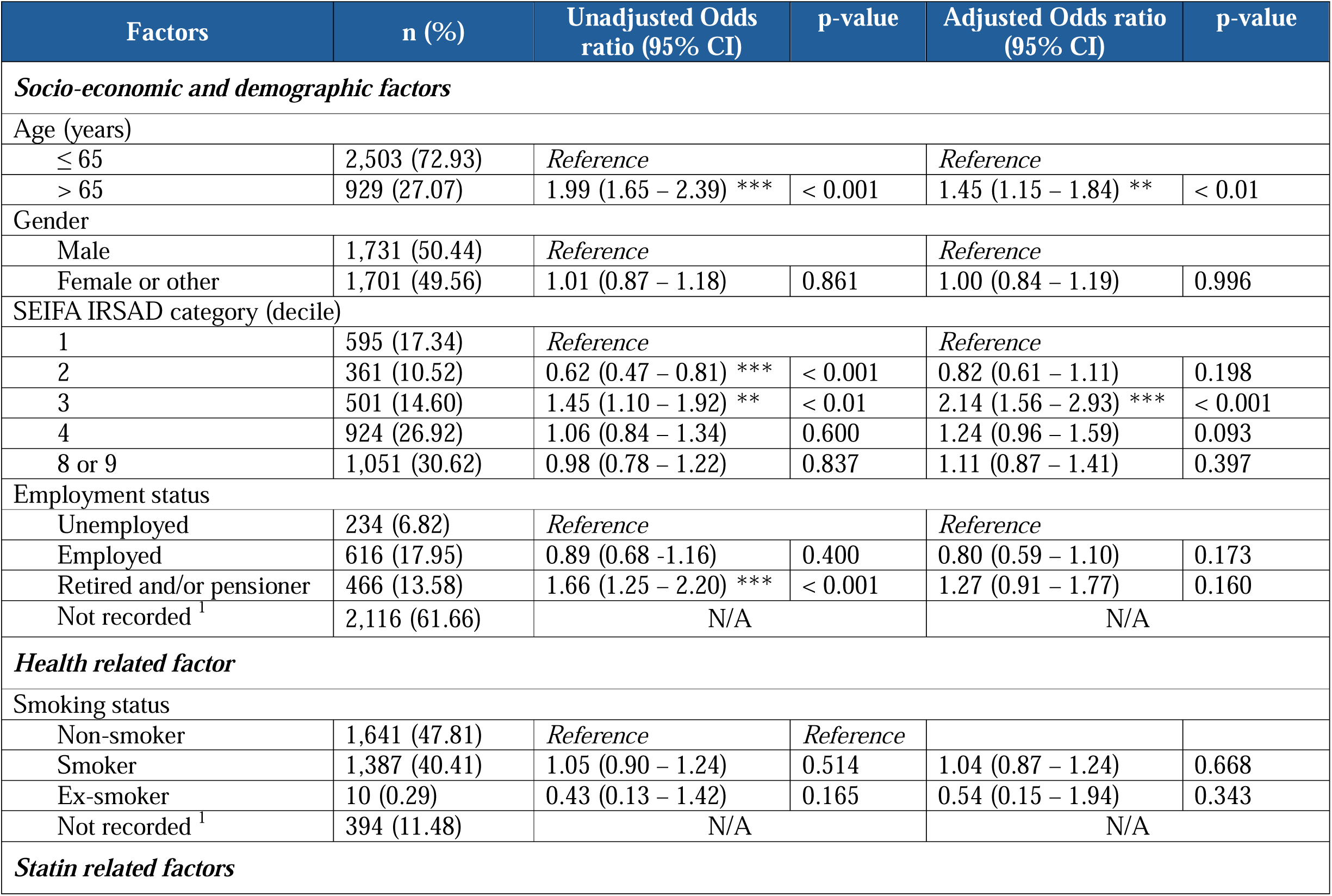
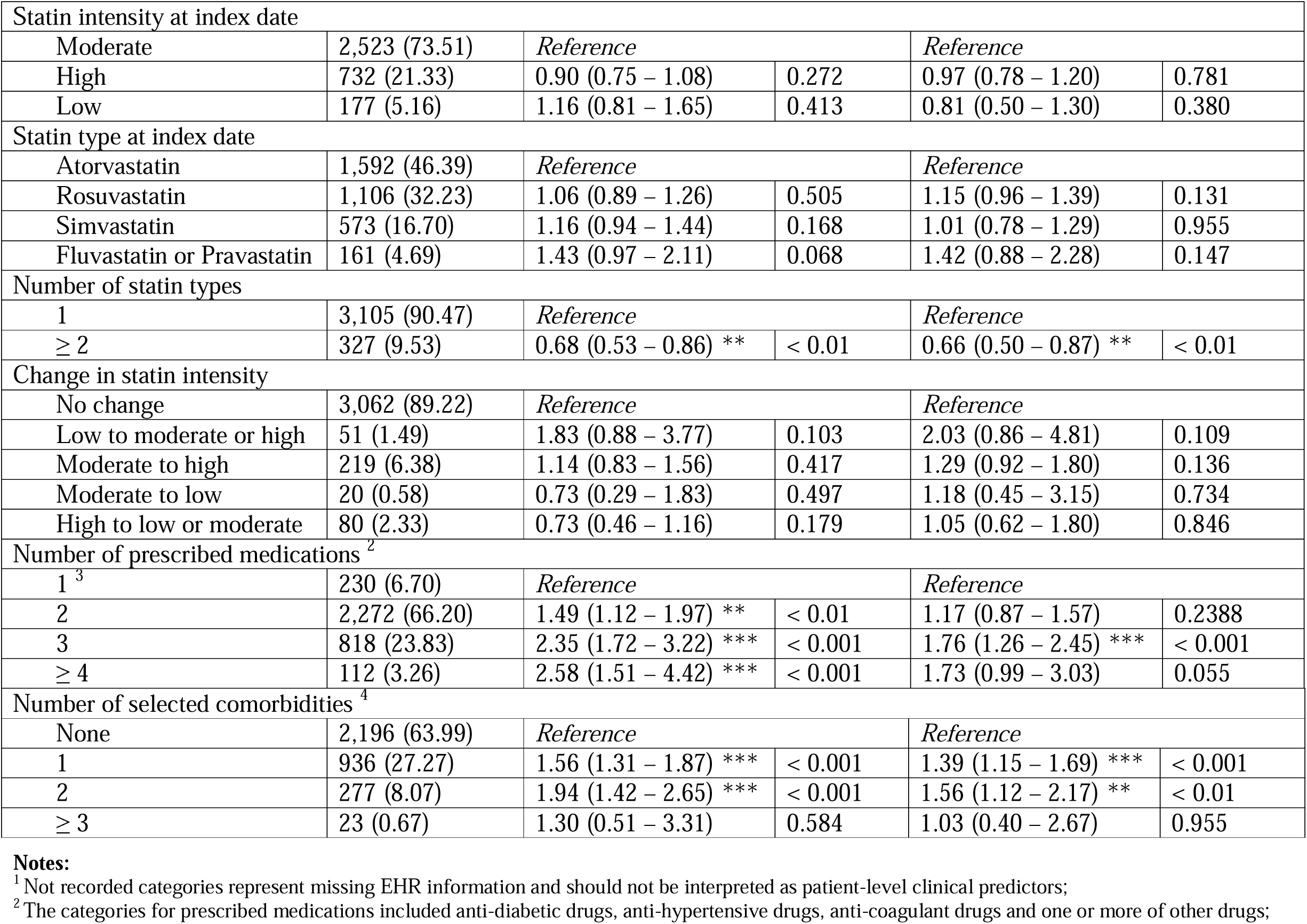
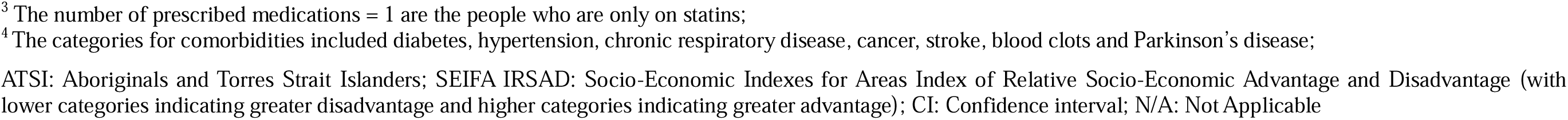
Univariate and multivariate logistic regression analysis for factors associated with statin adherence (*N*_=_3,432)

Lower odds of statin adherence were observed among patients prescribed two or more different statin types during the observation period (AOR 0.7, 95% CI 0.5 – 0.9), compared with the reference category.

### 3.5 Predictors of statin adherence in the final model

The logistic regression of the overall model met all key assumptions. The components of the overall regression model and their explanations are shown in **Supplement 4**. The overall model showed adequate fit (Hosmer-Lemeshow test: p = 0.20) and modest discriminative ability (AUC = 0.64). **Supplement 5** shows the details on the assumptions of regression of the overall model.

A reduced model was derived through backward stepwise selection based on AIC **(Table 3)**. Compared with the overall model, the final model had a lower AIC, while the likelihood ratio test showed no significant loss of model fit following the exclusion of variables **(Supplement 6)**. Discrimination was modest for both models, and the difference between the AUCs was statistically significant, although the magnitude of this difference was small **(Supplement 7)**.

**Table 3:** Predictors of statin adherence identified in the final multivariate logistic regression model.

| Factors | n (%) | Adjusted Odds ratio<br>(95% CI) | p-value |
| --- | --- | --- | --- |
| ***Socio-economic and demographic factors*** |  |  |  |
| Age (years) |  |  |  |
| ≤ 65 | 2,503 (72.93) | Reference |  |
| > 65 | 929 (27.07) | 1.42 (1.13 – 1.80) ** | < 0.01 |
| SEIFA IRSAD category (decile) |  |  |  |
| 1 | 595 (17.34) | Reference |  |
| 2 | 361 (10.52) | 0.80 (0.60 – 1.08) | 0.147 |
| 3 | 501 (14.60) | 2.11 (1.55 – 2.87) *** | < 0.001 |
| 4 | 924 (26.92) | 1.24 (0.97 – 1.59) | 0.081 |
| 8 or 9 | 1,051 (30.62) | 1.11 (0.88 – 1.41) | 0.6386 |
| Employment status |  |  |  |
| Unemployed | 234 (6.82) | Reference |  |
| Employed | 616 (17.95) | 0.82 (0.61 – 1.09) | 0.168 |
| Retired and/or pensioner | 466 (13.58) | 1.28 (0.94 – 1.75) | 0.121 |
| Not recorded <sup>1</sup> | 2,116 (61.66) | N/A |  |
| ***Statin related factors*** |  |  |  |
| Number of statin types |  |  |  |
| 1 | 3,105 (90.47) | Reference |  |
| ≥ 2 | 327 (9.53) | 0.71 (0.55 – 0.90) ** | < 0.01 |
| Number of prescribed medications <sup>2</sup> |  |  |  |
| 1 <sup>3</sup> | 230 (6.70) | Reference |  |
| 2 | 2,272 (66.20) | 1.18 (0.88 – 1.58) | 0.261 |
| 3 | 818 (23.83) | 1.75 (1.25 – 2.43) *** | < 0.001 |
| ≥ 4 | 112 (3.26) | 1.71 (0.98 – 2.99) * | 0.060 |
| Number of selected comorbidities <sup>4</sup> |  |  |  |
| None | 2,196 (63.99) | *Reference* |  |
| 1 | 936 (27.27) | 1.38 (1.14 – 1.68) *** | < 0.001 |
| 2 | 277 (8.07) | 1.54 (1.10 – 2.15) * | < 0.05 |
| 3 | 23 (0.67) | 1.01 (0.39 – 2.62) | 0.983 |

**Notes:**
<sup>1</sup> Not recorded categories represent missing EHR information and should not be interpreted as patient-level clinical predictors;
<sup>2</sup> The categories for prescribed medications included anti-diabetic drugs, anti-hypertensive drugs, anti-coagulant drugs and one or more of other drugs;
<sup>3</sup> The number of prescribed medications = 1 are the people who are only on statins;
<sup>4</sup> The categories for comorbidities included diabetes, hypertension, chronic respiratory disease, cancer, stroke, blood clots and Parkinson's disease;
- Variables not included in the final model for sensitivity analysis;
\* p < 0.05;
\*\* p < 0.01;
\*\*\* p < 0.001;
ATSI: Aboriginals and Torres Strait Islanders; SEIFA IRSAD: Socio-Economic Indexes for Areas Index of Relative Socio-Economic Advantage and Disadvantage (with lower categories indicating greater disadvantage and higher categories indicating greater advantage); CI: Confidence interval; N/A: Not Applicable

## 4. Discussion

In this study, we found that the mean PDC was 91.6% (±22.2%), and that 72.0% of patients were adherent with PDC ≥ 80% during the first two years after the index date. Although the mean PDC value aligned with findings from some international studies, it was higher than what is reported in previous Australian studies (47, 48, 57). In contrast, the 72% adherence rate was comparable to previous Australian studies (47, 48). An important difference is the source of medication data used to measure adherence. Previous Australian studies primarily used medication dispensing records from sources such as Pharmaceutical Benefits Scheme (PBS) and Health Insurance Bureau (HIB), which indicate that a prescribed medication was dispensed to the patient (47, 48). In comparison, the ePBRN data contain GP prescription records, which indicate that a prescription was issued but do not confirm that the medication was dispensed or taken. As such, adherence may have been overestimated in this study as it used electronic prescription records, in which overlapping prescriptions, dose changes or repeated prescriptions could be reflected in the estimated medication supply, thereby increasing the estimated adherence (58, 59). In addition, this study used a modified PDC in which the denominator extended from the index date to the last recorded visit within the two years observation period, rather than using a fixed 730 days denominator. This approach aligned the denominator with the period over which medication coverage was estimated using the available prescribing information. However, the shorter denominator for patients whose last recorded visit occurred before day 730 may have produced higher PDC estimates than a conventional calculation using a fixed two years denominator. Differences in both the medication data source and the PDC calculation should therefore be considered when comparing these findings with previous Australian studies. Moreover, South West Sydney includes a culturally and linguistically diverse (CALD) population across different socioeconomic groups, with documented health inequities, variable access to care, and a high prevalence of chronic diseases (41). In such settings, adherence to long-term lipid-management regimens or other statin-related therapies among the CALD population may differ from the population in other Australian studies as adherence may be influenced by factors such as health literacy, language barriers, access to primary care, affordability, and perceived side effects (42, 60).

This study found that sociodemographic factors such as age and SEIFA index were positively associated with statin adherence. Patients aged > 65 years showed about 45% higher odds of adherence compared to those who were ≤ 65 years, which aligns with the findings from other studies and systematic reviews (7, 12). Age-related adherence patterns have been noted across statins and other chronic medicines, but there is substantial variation based on setting, comorbidity burden, and access to care (61, 62). Previous studies have suggested that greater awareness of cardiovascular risk and the perceived importance of preventive treatment may contribute to higher adherence among some older patients (13, 14). On the other hand, this study found that patients from a relatively disadvantaged area (SEIFA IRSAD category 3) had about 110% higher odds of adherence compared to those from more disadvantaged areas (SEIFA IRSAD category 1). This finding suggests that the relationship between area level socioeconomic disadvantage and statin adherence may be complex and may not follow a consistent gradient across socioeconomic categories. In Australia, socioeconomic advantage and disadvantage as well as living in remote areas are often linked with lower adherence because of financial strain, health literacy, access barriers, and other social determinants (50, 63). These relationships may also vary across populations, including the diverse population of South Western Sydney, where health literacy and comorbidity burden may differ between patient groups (21, 50, 63).

The number of prescribed medications and the number of selected comorbidities were also positively associated with statin adherence. Previous studies have reported varying relationships between medication use, comorbidity, and adherence, with greater treatment complexity sometimes associated with poorer adherence because of factors such as competing treatment priorities and more complex medication regimens (64, 65, 66, 67, 68). However, direct comparison with these studies should be made cautiously. The medication measure used in this study represented the number of prescribed medications recorded in the EHR and was not a conventional measure of polypharmacy. Similarly, the comorbidity count was restricted to the selected conditions included in the analysis and did not represent patients’ complete comorbidity burden. The observed associations should therefore be interpreted specifically in relation to these measures rather than as evidence that greater overall medication or comorbidity burden improves statin adherence.

This study also found that patients prescribed more than one statin type had lower odds of adherence. Previous studies have reported that switching between statins may occur alongside difficulties maintaining treatment, including suspected adverse effects or treatment changes (69, 70). However, the temporal relationship between statin switching and adherence could not be established in this study. Missing information was also observed for smoking status, employment status, and ethnicity. Incomplete documentation of patient characteristics is a recognized limitation of routinely collected EHR data and may affect the interpretation of findings derived from these records (71, 72, 73, 74, 75, 76). To address missing employment and smoking information, MICE was performed, allowing patients with incomplete records for these variables to be retained in the regression analysis. However, this approach relied on the assumption that the missing data were missing at random, conditional on the observed information included in the imputation model. As this assumption could not be verified using the available EHR data, some uncertainty remains regarding the imputed values and their potential influence on the estimated associations. This is particularly relevant for employment status, given the substantial proportion of missing information. Therefore, the associations involving employment and smoking status should be interpreted cautiously.

Similar to findings from a systematic review of 102 conceptual frameworks on medication adherence, most of the factors assessed in this study were patient related, with additional factors related to medication, health conditions, and socioeconomic circumstances (77). However, other factors identified in the wider literature, such as patient beliefs about treatment, perceived benefits, health literacy, social support, and practical barriers to accessing care, could not be assessed in this study because these are not routinely captured in the ePBRN data (78). A qualitative study of patients and healthcare professionals also identified several barriers to the use of recommended medical therapy, including limited awareness of guidelines, concerns about their applicability to patients with complex conditions, treatment costs, frequent referrals, limited collaboration between healthcare professionals, short consultation times, and limited patient knowledge about medications (79). These findings highlight that medication adherence is influenced not only by patient and clinical characteristics, but also by healthcare providers and the wider healthcare system. These additional factors were not available in the ePBRN data and may help explain aspects of statin adherence that could not be accounted for by the predictors examined in this study.

### Limitations

This study used the ePBRN Linked Dataset from general practices, which contains prescription records rather than pharmacy dispensing records from beneficiary schemes such as PBS. Therefore, the available data indicate that a medication was prescribed but cannot confirm whether it was dispensed or taken by the patient. Replacement prescriptions for lost or misplaced scripts, overlapping prescriptions, early refills, or changes in treatment may appear as additional medication supply and could affect the estimation of statin adherence. Prescriptions issued by healthcare providers outside participating ePBRN practices may also not have been captured, resulting in incomplete information on patients’ statin use during the observation period. Prior statin exposure was assessed using the complete prescribing history available for each patient within the ePBRN records. However, prescriptions received outside participating practices before entry into the study could not be identified. As a result, the assigned index date reflects the earliest statin prescription observable within this dataset rather than necessarily indicating initiation of statin therapy for the first time. The modified PDC used in this study also differs from the conventional PDC calculation. The denominator was based on the period between the index date and the last recorded visit within the two years observation period rather than a fixed two years period. This approach was used because medication coverage could not be established for an exact two years period for every patient using the available prescription data, and to avoid estimating medication coverage beyond the predefined observation period. Although all included patients had at least two years of available EHR follow-up, the timing of the last recorded GP visit within the predefined two years observation period varied between patients and could occur before day 730. The modified PDC denominator therefore extended from the index date to the last recorded visit within this period. Medication coverage for the final interval was estimated using prescription information from the second last recorded visit, based on the assumption that the prescribed statin provided coverage until the subsequent visit. This approach allowed adherence to be estimated over the period for which medication use could reasonably be inferred from the available prescribing records, rather than assuming noncoverage for periods without a recorded encounter. However, as the ePBRN data contain prescribing rather than dispensing or medication taking information, actual medication possession during this interval could not be confirmed. The modified approach may therefore produce different adherence estimates from a conventional PDC calculated using a fixed denominator of 730 days.

The measure of medication use and comorbidity were limited to the information available and predefined for this analysis. The medication count reflected the number of prescribed medications recorded in the EHR and should not be interpreted as a conventional measure of polypharmacy. Similarly, the comorbidity count included only the selected conditions examined in this study and did not represent the full range of conditions that patients may have had. Consequently, medication use and overall comorbidity burden may have been underestimated for some patients. This may have influenced the observed associations with statin adherence, while medications and health conditions not adequately captured by these measures may have contributed to residual confounding.

Smoking and employment status were incompletely recorded in the ePBRN dataset. To address this, MICE was performed rather than treating missing information as a separate “not recorded” category in the regression analysis. This approach allowed patients with incomplete information for these variables to be retained in the analysis and accounted for uncertainty associated with the missing values through the generation of multiple imputed datasets. However, MICE relied on the assumption that the missing data were missing at random, conditional on the observed information included in the imputation model. This assumption could not be verified using the available EHR data. Given the substantial proportion of missing employment information, the imputed values may not fully represent the underlying employment characteristics of the study population, particularly if missingness was associated with factors not captured in the dataset. Consequently, the estimated associations involving employment and smoking status should be interpreted cautiously, as they may be influenced by uncertainty in the imputation process and unmeasured factors associated with missing documentation.

Finally, the findings were derived from a single EHR data source covering general practices in South Western Sydney, and may not be directly generalizable to populations or healthcare settings with different demographic, clinical, or prescribing characteristics. The exclusion of patients who experienced a CVD event before the index date or during the observation period may also have introduced selection bias by removing patients at higher short-term cardiovascular risk. This further limits the generalizability of the findings to the broader population of patients prescribed statins in primary care. In addition, the overall regression model demonstrated modest discriminative ability, suggesting that the variables available in the ePBRN dataset captured only part of the variation in statin adherence. Other patient, treatment, and healthcare factors not available in the dataset may also contribute to adherence.

## 5. Conclusion

This study found that the mean PDC was 91.6% (±22.2%), with 72.0% of patients classified as adherent (PDC ≥ 80%) during the first two years after the index date. Higher odds of statin adherence were associated with older age, SEIFA IRSAD category 3, the number of prescribed medications, and the number of selected comorbidities, while patients prescribed two statin types had lower odds of adherence. These findings highlight the importance of considering sociodemographic and treatment related characteristics when examining statin adherence in primary care. However, the medication and comorbidity measures were limited to the information captured in the EHR, and the modified PDC was based on prescribing rather than dispensing data. The findings may therefore not be directly generalizable to other populations or settings. Further research using independent datasets and additional patient and healthcare factors is needed to validate these associations and inform the development of targeted strategies to support statin adherence in primary care.

## Ethics statement

This study and the use of data was approved by the UNSW Human Research Ethics Advisory Panel (HC230066; 23 June 2023).

## Data Availability

The corresponding author will make the data available upon request.

